# Validation of Utstein-Based score to predict Return Of Spontaneous Circulation (UB-ROSC) in patients with out-of-hospital cardiac arrest

**DOI:** 10.1101/2023.07.05.23292272

**Authors:** Maria Luce Caputo, Enrico Baldi, Roman Burkart, André Wilmes, Ruggero Cresta, Claudio Benvenuti, Roberto Cianella, Roberto Primi, Alessia Currao, Sara Bendotti, Sara Compagnoni, Francesca Romana Gentile, Luciano Anselmi, Simone Savastano, Catherine Klersy, Angelo Auricchio

## Abstract

**Background:** Prediction of probability of return of spontaneous circulation (ROSC) during out-of-hospital cardiac arrest (OHCA) is one of the biggest challenge in resuscitation science. The Utstein Based-ROSC (UB-ROSC) score has been developed to predict ROSC in OHCA’s victims. Aim of the study was to validate UB-ROSC score using two large Utstein-based OHCA registries: the SWiss REgistry of Cardiac Arrest (SWISSRECA) and the Lombardia Cardiac Arrest Registry (Lombardia CARe), northern Italy.

**Methods:** Consecutive OHCA of any etiology occurring between January 1^st^, 2019 and December 31^st^ and registered in 2 large national registries were included in a prospectively designed validation study. For model validation, a model area under the ROC curve (AUC ROC) for discrimination, using 10-fold cross-validation was computed. The score was plotted against the survival to hospital admission.

**Results:** 14,715 patients were included in the study. A sustained ROSC was obtained in 3,151 patients (21%). Overall, UB-ROSC model showed a good discrimination (AUC 0.72, 95% CI 0.71-0.73). Shape of risk predicted by the model was almost linear and the model resulted well calibrated. In the low likelihood subgroup of UB-ROSC, only 9% of patients achieved a ROSC. This proportion raised to 35% for UB-ROSC score between −18 and 12 (OR 5.3, 95% CI 2.9-9.4, P<0.001). Finally, in 85% of patients with UB-ROSC values of more than or equal to 13 a ROSC was obtained (OR 52.1, 95%CI 15.6-173.0, P <0.001).

**Conclusions:** UB-ROSC score may represent a reliable tool to predict ROSC probability. Its application may help the decision-making process providing a realistic stratification of probability of ROSC achievement.

**Clinical perspectives:** **What is new?**

- UB-ROSC is the unique Utstein-based score designed to help EMS staff to stratify patient’s probability of ROSC before treating the patient.
- In this validation study, UB-ROSC score was verified in a cohort of more than 14,000 OHCA and confirmed a very high power in discrimination of ROSC probability
- The score is very user-friendly and may be considered a helpful tool for EMS in the decision-making process when approaching OHCA victims.

**What are the clinical implications?**

- Sudden cardiac death is the leading causes of death worldwide and substantially contribute to loss of health and excess health system costs.
- Development and validation of models to stratify probability of survival are increasingly necessary in the decision-making process, particularly in a pre-hospital setting, to offer a realistic expectation of survival and eventually to terminate resuscitation attempts.

## Introduction

Out-of-hospital cardiac arrest (OHCA) is the most frequent cause of death in the industrialized countries (1,2). OHCA outcome is influenced by several independent variables related to patient’s characteristics, e.g. age, comorbidity, and to circumstances of the event (etiology, witnessed or not, public or private location) (3) as well as by modifiable factors, including bystanders and intervention time of emergency medical services (EMS) (4,5). As survival, also the probability to return of spontaneous circulation (ROSC) significantly varies across geographies (3,6).

Prediction of sustained ROSC is an important goal in resuscitation science. Its assessment is highly valued by both medical personnel, and family members as it may allow a reasonable expectation of immediate outcome as well as to provide a guidance for termination-of-resuscitation rules beyond those currently available. Among other scores, the Utstein Based-ROSC (UB-ROSC) score was recently proposed as a multiparametric operative model developed using pre-hospital independent variables, collected according to Utstein recommendations (7). It allows to predict probability of sustained ROSC leading to hospital admission with a good sensitivity and specificity (7). The score was developed and validated in two large regional registries, the TIRECA (TIcino REgstry of Cardiac Arrest) in the Swiss Canton Ticino and the Pavia CARe (Pavia Cardiac Arrest Registry) in the Italian province of Pavia. The score can be calculated using a mobile application (http://www.sanmatteo.org/site/home/ub-rosc-score.html) (8–11). To date, the UB-ROSC score has not been never validated in a larger external cohort of OHCAs.

The aim of our study was to validate the UB-ROSC using OHCAs collected in two large Utstein-based registries in two different countries: the SWISSRECA (Swiss Registry of Cardiac Arrest) and the Lombardia CARe (Lombardia Cardiac Arrest Registry).

## Methods

### Participants

All consecutive patients who suffered an OHCA of any etiology between 1st of January 2019 and 31st of December of 2021 in Lombardia, northern Italy, and in Switzerland were included in the study. Patients declared dead before ambulance arrival, with a “do not resuscitate” order, with incomplete or unknown data were excluded from further analysis.

### Study design and setting

This is a prospectively designed study to validate a multiparametric operative score (UB-ROSC score) capable to predict the probability of survival to hospital admission (sustained ROSC) by using consecutive OHCA cases already available in 2 large national registries. UB-ROSC score development was described elsewhere (7). Lombardia region and Switzerland both have a prospectively designed registry of cardiac arrest. Both registries follow the Utstein recommendations for data collection (12,13). The two registries are periodically reviewed for quality assessment by an internal commission, and were approved by the local ethical committee. The Lombardia CARe enrolls all the OHCA cases occurring in the Province of Pavia since January 1, 2015 and in the provinces of Pavia, Lodi, Cremona, and Mantua since January 1, 2019. All the data are collected following Utstein 2014 recommendations (11). The registry was approved by the Ethical Committee of the Fondazione IRCSS Policlinico San Matteo (proc. 20140028219) and by all others who were territorially involved. An informed consent form was signed by all the patients discharged alive. SWISSRECA is a national cardiac arrest registry set up by the Interassociation for Rescue Services (IVR-IAS) at the end of 2018, which collects OHCAs of every etiology occurring in the whole of the Switzerland. SWISSRECA is approved by the national Ethical committee (Swissethics-ID-2016-01844) and IVR-IAS is responsible for its maintenance.

### EMS and resuscitation network in Lombardia Region, Northern Italy

The total area covered by the Lombardia CARe registry was 7,863 km^2^ in 2019 and increased to 15,125 km^2^ in 2021. Details of registry development and corresponding changes over time in provinces included in the registry are reported in table 1 of supplemental materials. Each province has several rural regions and a few urban areas for a total population covered of 1,547,333 inhabitants in 2019, increased to 4,274,296 inhabitants in 2021.

**Table 1.** Variables and corresponding scores for UB-ROSC calculation. Witness status and cardiopulmonary resuscitation (CPR) before ambulance arrival were combined as follows: no wit/no CPR: bystander witnessed: no, CPR before ambulance arrival: no; no wit/yes CPR: bystander witnessed: no, CPR before ambulance arrival: yes; yes wit/no CPR: bystander witnessed: yes, CPR before ambulance arrival: no; yes wit/no CPR: bystander witnessed: yes, CPR before ambulance arrival: yes; EMS: emergency medical service.

| UB-ROSC variables | SCORE COMPONENT |
| --- | --- |
| **Sex** |  |
| Female | 0 |
| Male | -3 |
| **Age** |  |
| <80 | 0 |
| ≥80 | -9 |
| **Aetiology** |  |
| Cardiac | 0 |
| Trauma | -3 |
| Drowning | 1 |
| Respiratory | 19 |
| Other non-cardiac | 0 |
| **Location** |  |
| At home | 0 |
| Nursing home | -7 |
| Workplace | 6 |
| School | 0 |
| Street | 4 |
| Public building | 5 |
| Sport | 7 |
| **Bystander and CPR** |  |
| No witnessed, no CPR | 0 |
| No witnessed, yes CPR | -5 |
| Witnessed, no CPR | 2 |
| Witnessed, yes CPR | 4 |
| EMS witnessed | 13 |
| **Rhythm** |  |
| Not shockable | 0 |
| Shockable | 21 |
| **Time to EMS arrival** |  |
| ≤ 10 min | 0 |
| 11-15 min | -4 |
| ≥15 min | -7 |
| **Constant** | -16 |

The EMS dispatch center coordinates ambulances staffed with basic life support and defibrillation (BLS-D)-trained personnel, and advanced life support (ALS)-trained staffed vehicles (a physician and a specialized nurse or a specialized nurse only). The specialized nurse, if alone in the ALS-staffed vehicle, applies the same ALS protocol, using supraglottic devices instead of tracheal intubation. Five helicopters with a physician and a specialized nurse on board also serve the entire region of Lombardy and another three can intervene from other neighboring regions. In the case of suspected OHCA, the EMS dispatcher activates one to three emergency vehicles (which may include a helicopter) with at least one physician and assists the calling bystander during chest compressions (telephone CPR). The decisions about the attempt and the duration of resuscitation are left to the physician whilst BLS-D-trained personnel are instructed to start resuscitation unless clear signs of death are present (rigor mortis, hypostasis, and injuries not compatible with life).

### EMS and resuscitation network in Switzerland

In Switzerland (8.57-million inhabitants over a territory of 41,285 Km^2^, divided in 26 Cantons), OHCAs are managed by local EMS with a two-tiered response system, coordinated via a regional (cantonal) dispatch center. The first tier consists of paramedics who can provide advanced life support, the second tier is made up of teams (ambulance or helicopter) with an emergency physician, alerted if required. Paramedics are instructed to initiate resuscitation unless clear signs of death are present or in case of a Do Not Attempt Resuscitation order. The decision to stop resuscitation and the death declaration are based on the physician’s clinical judgment. The Swiss territory encompasses several rural areas (mountains and valleys) and few relatively small urban areas. In all cantons, there is a network of first responders (FR), made up of off-duty EMS personnel, fire-fighters, police and laypeople trained in CPR, who are alerted via a mobile application and can provide basic life support and use an automatic external defibrillator (AED).

### UB-ROSC score variables and outcome of the model

UB-ROSC score model development was described before (7). Briefly, variables that may be determinants of survival at hospital admission were identified among Utstein variables immediately available at EMS arrival on the scene. Age, sex, aetiology, location, witnessed OHCA, bystander CPR, time of EMS arrival and shockable rhythm were included in the model (7). The coefficients estimated from the model, multiplied by 10 and rounded to the closest integer were used to compute a prediction score. All variables included in the model and corresponding scores for UB-ROSC calculation were reported in table 1. Survival to hospital admission was defined as patient with ROSC sustained until arrival at the emergency department and transfer of care to medical staff at the receiving hospital. This definition corresponds to the Utstein recommendations’ core outcome of “Survived event” (13). Probability of survival at hospital admission corresponding to different values of UB-ROSC score were reported in table 2 of Supplemental Material.

**Table 2.** Comparison between out-of-hospital cardiac arrest presentation and outcome in the two registries. Witness status and cardiopulmonary resuscitation (CPR) before ambulance arrival were combined as follows: no wit/no CPR: bystander witnessed: no, CPR before ambulance arrival: no; no wit/yes CPR: bystander witnessed: no, CPR before ambulance arrival: yes; yes wit/no CPR: bystander witnessed: yes, CPR before ambulance arrival: no; yes wit/no CPR: bystander witnessed: yes, CPR before ambulance arrival: yes; EMS: emergency medical service. ROSC: return of spontaneous circulation.

|  | **Lombardia Care**<br>**N=7663** | **SWISSRECA**<br>**N=7052** | **P value** |
| --- | --- | --- | --- |
| **Sex, n (%)** |  |  | <0.001 |
| Female | 3071 (40) | 2050 (29) |  |
| Male | 4544 (60) | 5002 (71) |  |
| **Age, median (IQR)** | 78 (66-86) | 70 (58-80) | <0.001 |
| **Medical etiology, n (%)** |  |  | 0.807 |
| Yes | 7094 (92) | 6305 (89) |  |
| No | 564 (8) | 747 (11) |  |
| **Location, n (%)** |  |  |  |
| Public | 1328 (17) | 2629 (37) | <0.001 |
| Private | 6335 (83) | 4423 (63) |  |
| **Shockable rhythm, n (%)** |  |  | <0.001 |
| Yes | 1051 (14) | 2201 (31) |  |
| No | 6510 (86) | 4851 (69) |  |
| **Combined Witness/CPR**<br>**before EMS arrival, n (%)** |  |  | <0.001 |
| No wit/no CPR | 1862 (25) | 709 (10) |  |
| No wit/yes CPR | 860 (11) | 1674 (24) |  |
| Yes wit/no CPR | 1766 (24) | 1002 (14) |  |
| Yes wit/ yes CPR | 2073 (28) | 2668 (38) |  |
| EMS witnessed | 906 (12) | 999 (14) |  |
| **Time to EMS arrival, min,**<br>**median (IQR)** | 13 (10-16) | 10 (8-13) | <0.001 |
| **Total resuscitation time, min,**<br>**median (IQR)** | 26 (16-40) | 23 (15-33) | <0.001 |
| **ROSC, n (%)** | 1090 (15) | 2061 (31) | <0.001 |

### Statistical analysis

All analyses were performed using the Stata software (release 17, College Station, TX, USA). A 2-sided p<0.05 was considered statistically significant. Continuous data are reported as mean and standard deviation or median and quartiles if skewed. Categorical data are reported as counts and percent. Data were compared between groups of patients (by national territory and by ROSC groups) with the Mann Whitney U test and the Fisher exact test, respectively. The UB-ROSC score was computed for each patient according to the original paper (7) and categorized in the 3 subgroups of low, medium and high likelihood of ROSC according to the published cut-offs of UB-ROSC. (≤-19; −18 to 12; ≥13). The latter variable was included in a logistic model for ROSC as the independent variable; Huber-White robust standard errors were computed to account for intra-region correlation. We derived the odds ratios (OR) and 95% confidence intervals (95%CI). To assess the performance of the UB-ROSC score in this new cohort, we assessed bot discrimination and calibration. For discrimination we computed the cross-validated model area under the Receiver Operating Characteristic-ROC curve and 95%CI. Sensitivity and specificity of the model were also computed. For this purpose, patients were classified as ROSC if the predicted probability was equal to or above 0.5. To assess calibration, we plotted the observed and predicted probabilities of ROSC as a calibration belt. Model goodness of fit was assessed with the Pearson test and was always satisfied.

## Results

Overall, 22,794 OHCA cases were included in the two registries. 8,079 (35%) were declared dead before ambulance arrival or had a “do not resuscitate” order, leaving. 14,715 patients for further analysis. Demographic and clinical characteristics of OHCAs included in the two national registries are reported in table 2. Swiss patients were in median 8-year younger than Italian ones (70 years old, IQR 58-80 versus 78 years old, IQR 66-86, respectively, P<0.001, table 2). Moreover, Swiss OHCAs occurred more often in public location (37% versus 17%, p<0.001, table 2) and had a first shockable rhythm (31% versus 14%, p<0.001).

### Observed ROSC and UB-ROSC validation

A ROSC was obtained in 3,151 patients (21%). The overall observed ROSC rate was higher in SWISSRECA than in Lombardia CARe (31% versus 15%, p<0.001, table 2). UB-ROSC model had a similar discrimination power with an area under the ROC curve (AUC) of 0.68 in SWISSRECA and of 0.73 in Lombardia CARe (figure 1). Model validation and calibration in the entire population is reported in figure 2. Overall, UB-ROSC model showed a good discrimination (AUC 0.72, 95% CI 0.71-0.73, figure 2, panel A). Model had 10% sensitivity and 99% specificity to predict ROSC, with a positive predictive value of 85% and a negative predictive value of 80%. Events correctly classified by the model were 80%. The shape of risk predicted by the model was almost linear and the model was very well calibrated, with a 10-fold cross validation AUC of ROC of 0.71, 95%CI 0.70-0-72, P 1.0. Curve of the shape of risk and internal calibration test is presented in figure 2, panel B and C. The probability to obtain ROSC, according to the 3 subgroups and corresponding ORs are reported in table 3 and Figure 3. For UB-ROSC score values of less than or equal to −19, only in 9% of patients a ROSC was achieved, increasing to 35% for UB-ROSC score between −-18 and 12 (OR 5.3, 95% CI 2.9-9.4, P<0.001). In 85% of patients with UB-ROSC values of more than or equal to 13 a ROSC was obtained (OR 52.1, 95%CI 15.6-173.0, P <0.001). Characteristics of OHCA presentation according to UB-ROSC score subgroups are reported in table 3. Patients with the lowest UB-ROSC values (≦-19) were significantly older (median age 80 years old, IQR 68-87), 52% unwitnessed, 83% occurred in private location and 99% of them had a no-shockable rhythm. In contrast, patients in the high likelihood subgroup of UB-ROSC (UB-ROSC score ≧ 13) were younger (median age 62 years old, IQR 53-71), 58% occurred in public location, and 99% of them were witnessed and with a first shockable rhythm, respectively.

**Figure 1.**
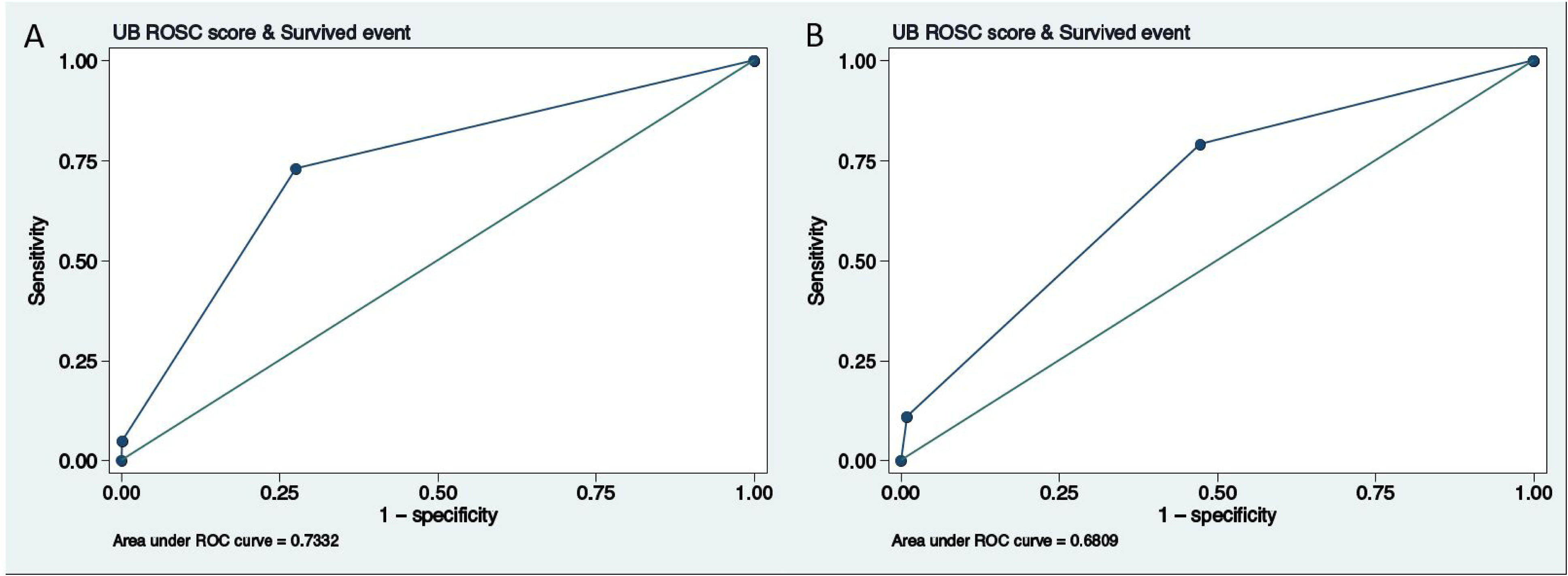
Receiver Operating Characteristic (ROC) curve of UB-ROSC score in Lombardia CARE (panel A) and in SWISSRECA population (panel B).

**Figure 2.**
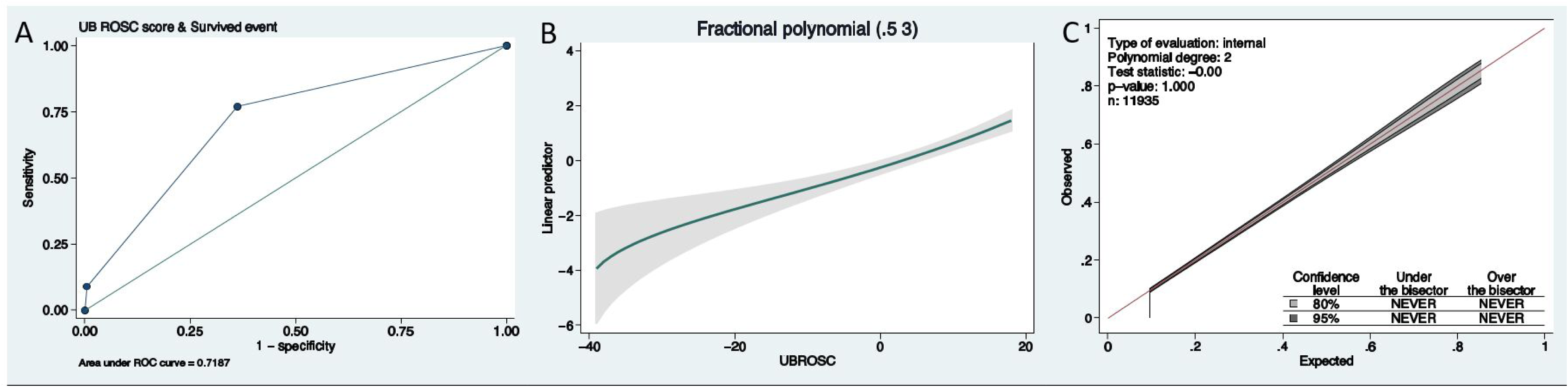
UB-ROSC model discrimination, shape of risk and calibration. **Panel A:** Receiver Operating Characteristic (ROC) curve of UB-ROSC score; **Panel B:** Shape of risk of the model. **Panel C:** Calibration curve of the model. The bisecting line corresponds to perfect calibration of the model (perfect agreement between observed ROSC and predicted ROSC. The line is entirely included in the shaded area corresponding to the 80% and 95% confidence intervals for the observed-predicted relationship, denoting that the model is well calibrated (there is neither over nor underestimation of mortality).

**Figure 3.**
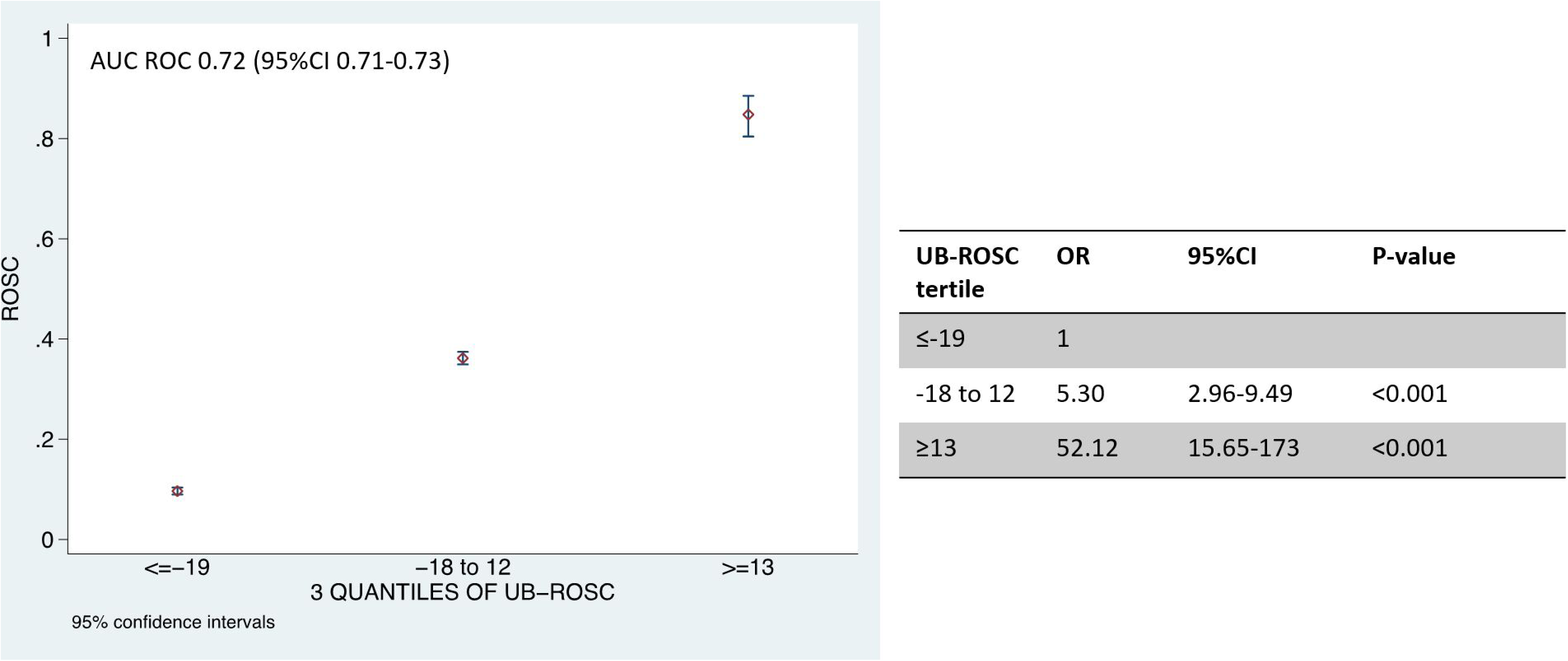
Cut-offs of UB-ROSC score and corresponding probability of ROSC.

**Table 3.**
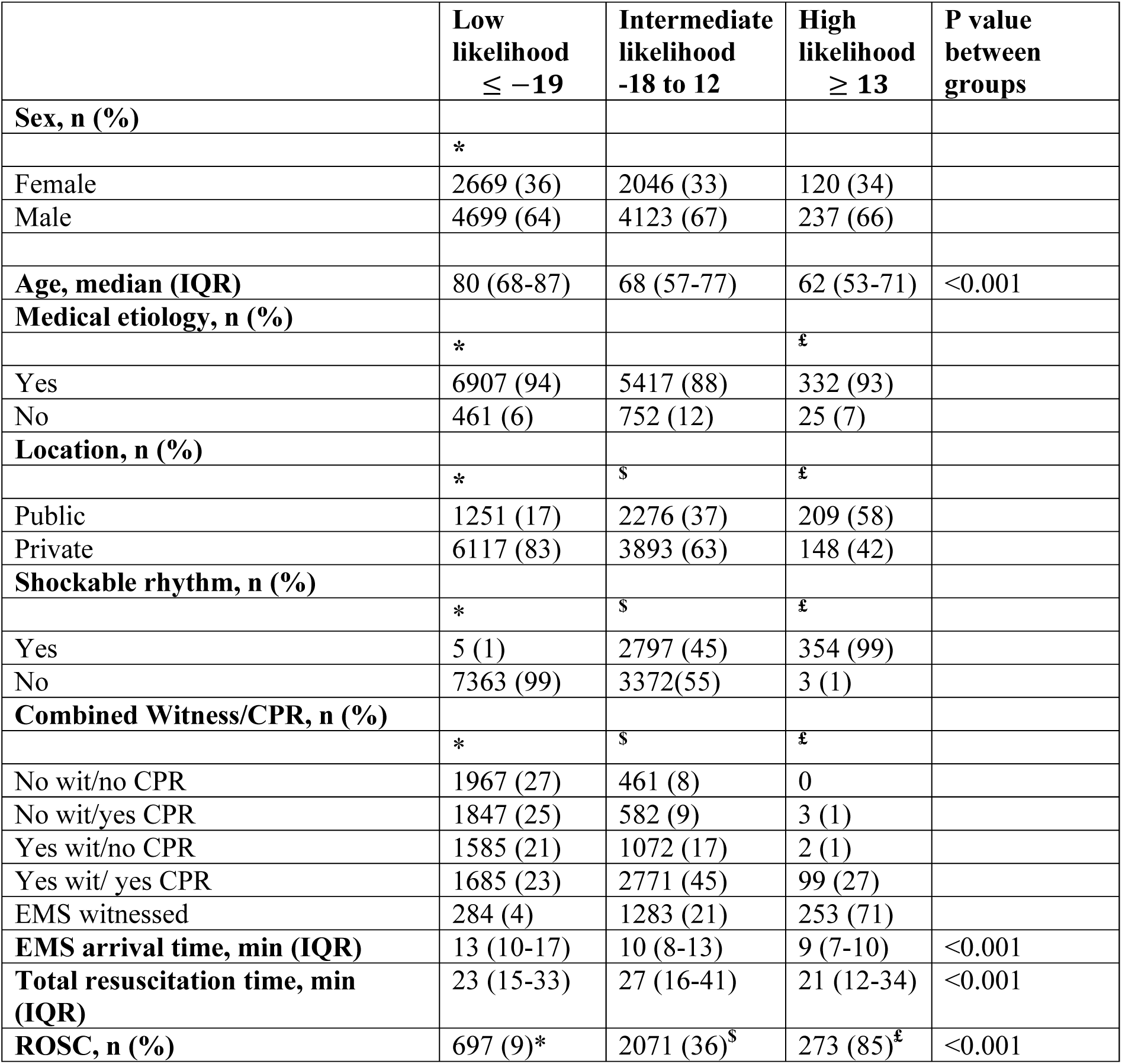
OHCA presentation characteristics according to UB-ROSC score likelihood groups. For categorical variables, Post-hoc comparisons p for significance was 0.017. * Significant p value between low and intermediate groups; ^$^ significant p value between intermediate and high groups; ^£^ significant p value between high and low groups. For continuous variables, p-value resulting from one-way analysis of variance by ranks (Kruskall-Wallis test) was reported. Witness status and cardiopulmonary resuscitation (CPR) before ambulance arrival were combined as follows: no wit/no CPR: bystander witnessed: no, CPR before ambulance arrival: no; no wit/yes CPR: bystander witnessed: no, CPR before ambulance arrival: yes; yes wit/no CPR: bystander witnessed: yes, CPR before ambulance arrival: no; yes wit/no CPR: bystander witnessed: yes, CPR before ambulance arrival: yes; EMS: emergency medical service. ROSC: return of spontaneous circulation.

## Discussion

UB-ROSC is an operational score for prediction of ROSC probability after an OHCA. It is designed to use Utstein variables and categories thus, allowing a very easy calculation of the score and of the corresponding probability of ROSC. For the first time, we showed in a large OHCA cohort that the UB-ROSC score had excellent discrimination capability and can be used in-field to possibly support resuscitation-related decisions such as considering termination of resuscitation manoeuvres, and to set realistic expectations about the likelihood of achieving sustained ROSC during resuscitation manoeuvres. This knowledge is important for paramedics, rescue teams and even more so for family members.

Similarly, ACLS score has been proposed as operative score (14) helping to stratify the probability of survival during ongoing resuscitation. The two models shared similarities in variables included but the performance of the two scores is not comparable, because ACLS score’s outcome was survival at discharge from hospital while UB-ROSC model predicts persistent ROSC (survival at hospital admission). Among various models that predict the likelihood of gaining ROSC, the ROSC after Cardiac Arrest (RACA) score developed with the German Resuscitation Registry (15) is one of the most validated in external populations. However, differently from UB-ROSC, the RACA score was not designed to be used as a prediction tool on the spot to facilitate resuscitation decisions. Instead, by providing a predicted ROSC rate, the score enables comparison of studies conducted in different communities and cohorts, serving as a ‘quality indicator’ of different EMS systems. External validation of RACA score yielded mixed results. (16–20) The discrepancies, although partially explained by different resuscitation practices over time, often imply the need for further adjustment of the score for individual communities. (18)

By using Utstein based variables, UB-ROSC score overcomes two major limitations of RACA score: first, the need to re-classify variables included in Utstein registries before calculate the score, and, then the potential bias of overestimation of ROSC by including also patients with transient ROSC. Moreover, when validated in an external population (17) the RACA score had a suboptimal calibration at the two extremes, i.e. in patients with the lowest or highest probability of ROSC. In contrast, UB-ROSC score confirmed a good discrimination power, with an AUC of ROC curve of 0.72, confirmed at 10-fold cross validation, and with an excellent calibration of the model even at the two extremes.

Swiss and Italian OHCA populations were significantly different in characteristics of presentation and in some aspects of the out-of-hospital management. As compared with Italian patients, Swiss patients were younger, more often had an OHCA in a public place and up to 50% of them received a CPR before ambulance arrival. However, even accounting for significant differences in the two enrolled population, UB-ROSC score confirmed a good power in prediction of ROSC probability with an excellent calibration of the model in both the two populations.

The increasing call for survival prediction models, eventually by including surrogates of quality of CPR, as End-tidal CO_2_ (21), or artificial intelligence software (22), derives from the emerging need to optimize available resources for pre-hospital ad in-hospital management of these patients. OHCA is a clinical event with huge repercussions on community in terms of costs, healthcare personnel utilization and psychological impact. Resuscitation prolongation and transportation to hospital of patients who have little possibility of survival has several negative consequences in terms of public health costs and noteworthy it represents an additional distress for family members. Starting from this consideration, several algorithms and rules have been proposed to help identifying those people with low chance of survival at discharge, in whom it could be reasonable to terminate resuscitation before hospital admission. UB-ROSC score cut-offs were very accurate in identification of patients with low probability of survival. Only 9% of patients in the lowest subgroup of UB ROSC score (≦-19) achieved a ROSC and, looking at their characteristics, they are almost all patients with no shockable rhythm, unwitnessed, and without CPR before ambulance arrival. Same characteristics have been included in different termination-of-resuscitation rules protocols. Accuracy of these protocols in predicting in-hospital mortality was assessed in large registries in Western countries and in Pan-Asian Resuscitation Outcomes Study (PAROS) registry. (23, 24) They identified a large proportion of patients who are candidates for termination of resuscitation following OHCA with very low rate of misclassifying eventual survivors (<0.1%). Even if patients fitting these rules had same characteristics of those with a low UB-ROSC score (unwitnessed OHCA, no shockable rhythm and no CPR initiated by bystander), evaluated outcome is substantially different. In a pre-hospital setting and when considering only pre-hospital variables, is probably more reasonable to evaluate ROSC probability instead of survival at discharge, being the latter affected also by in-hospital treatment. The same observation may apply also to ACLS score application, which evaluated survival at discharge. With this purpose, UB-ROSC score was designed to be calculated even before EMS team arrival on scene, in order to provide an estimation of ROSC likelihood before approaching the patient.

Tools like UB-ROSC may be useful also to individuate people with the theoretically highest chance of survival. Patients in the of UB-ROSC subgroup with highest likelihood of ROSC have a very favourable OHCA presentation (shockable rhythm, public place, bystander or EMS witnessed). Fast recognition of these patients may help to organize all available resources, eventually including early in-hospital hemodynamical support, and to personalize the resuscitation approach by taking in to account the real probability of survival.

Comparison between predicted and observed ROSC represents a unique opportunity for EMS staff to re-analyse and debrief most difficult cases. With this purpose, UB-ROSC is currently automatically calculated for each OHCA enrolled in SWISSRECA. This help a retrospective quality reassessment of those cases in which, despite a high UB-ROSC score, ROSC was not achieved.

### Limitations

This study has some limitations. First, the validation cohort is an European population and the two regions shared similarities in geographical context and in EMS organization. Moreover, the two population of the original validation cohort of UB-ROSC score are included in this validation cohort. Further studies are needed to assess UB-ROSC application in different geographical areas and with substantial differences in EMS organization (i.e. Asian countries). Last, UB-ROSC score is calculated using Utstein variables and categories. Potential application of the score in non-Utstein registries requires adjustment of variables and may determine unpredictable differences in the performance of the model that needs further assessment.

## Conclusions

UB-ROSC score is a valuable tool to predict ROSC probability after OHCA. Its validation in a large cohort of patients confirmed the good calibration of the model. Application of the score in a pre-hospital setting may help the decision-making process by providing a realistic stratification of probability of ROSC achievement.

## Data Availability

The dataset is available on request.

## Acknowledgments

We thank all the investigators of the Lombardia CARe and SWISSRECA Registry, the EMS personnel and all the people involved in the resuscitation network in our countries. EB and SS are part of the European Resuscitation Council (ERC)-Research NET. Lombardia CARe is partner of the ESCAPE-NET consortium. MLC, EB, FRG, SS and AA are part of the COST action PARQ.

The Lombardia CARe is a research project of the Fondazione IRCCS Policlinico San Matteo and partially supported by Fondazione Banca del Monte di Lombardia.

## Sources of Funding

### Disclosures

Authors have no conflict of interest to declare.

---

**Table 1_Supplemental material.** Provinces progressively included in Lombardia CARe registry since 2019.

**Table 2_Supplemental material.** Probabilities of survival at hospital admission with 95% CI and corresponding values of UB-ROSC score. (7)

## Notes

### Competing Interest Statement

The authors have declared no competing interest.

### Author Declarations

The Lombardia CARe registry was approved by the Ethical Committee of the Fondazione IRCSS Policlinico San Matteo (proc. 111 20140028219) and by all others who were territorially involved. SWISSRECA is approved by the national Ethical committee (Swissethics-ID-2016-01844) and IVR-IAS is responsible for its maintenance.

